# Relationships of total COVID-19 cases and deaths with ten demographic, economic and social indicators

**DOI:** 10.1101/2020.09.05.20188953

**Authors:** Dimitar Valev

## Abstract

The statistical relationships of total COVID-19 Cases and Deaths per million populations in 45 countries, where 85.8% of the world’s population lives with 10 demographic, economic and social indicators were studied. Data for 28 May 2020 were used in the main calculations. The relationship of Deaths per million population and total Cases per million population is very close and reaches correlation coefficient R = 0.926. It is interesting that the close correlations were found of Cases and Deaths per 1 million with a purely economic index like GDP PPP per capita, where R = 0.687 and R = 0.660, respectively. Even more close correlations were found of Cases and Deaths per 1 million with a composite index HDI, where the correlation coefficients reach 0.724 and 0.680, respectively. The main reason for these paradoxical results is the underestimation of pandemic restrictions in the form of masks, social distance and disinfection in most of these countries. Other indicators (excluding Gini index and Population Density) also show statistically significant correlations with Cases and Deaths per 1 million with correlation coefficients from 0.432 to 0.634. The statistical significance of the found correlations determined using Student’s t-test was p <0.0001. Surprisingly, there was no statistically significant correlation between Cases and Deaths with Population Density. To check whether there is a change in the correlations with the development of the pandemic, a statistical analysis was made for four different dates – 9 April, 28 May, 7 August and 30 November 2020. It was found that the correlation coefficients of COVID-19 cases and Deaths with the rest indicators decrease during the pandemic.

## I. INTRODUCTION

Coronavirus disease 2019 (COVID-19) is an infectious disease caused by severe acute respiratory syndrome coronavirus 2 (SARS-CoV-2) [1]. It was first identified in December 2019 in Wuhan, China, and has resulted in an on-going pandemic [2]. As of 24 November 2020, almost 60 million cases have been reported across 218 countries and territories, resulting in more than 1.4 million deaths. More than 41 million people have recovered [3]. Common symptoms include fever, cough, fatigue, shortness of breath, and loss of smell and taste. While the majority of cases result in mild symptoms, some progress to acute respiratory distress syndrome (ARDS) possibly precipitated by cytokine storm, multi-organ failure, septic shock, and blood clots. The virus is primarily spread between people during close contact, most often via small droplets produced by coughing, sneezing, and talking. According to the World Health Organization (WHO), there are neither vaccines nor specific antiviral treatments for COVID-19 [4]. Management involves the treatment of symptoms, supportive care, isolation, and experimental measures. The WHO recommends 1 - 2 meters of social distance.

Severe acute respiratory syndrome coronavirus 2 (SARS-CoV-2) is a novel severe acute respiratory syndrome coronavirus closely related to the original SARS-CoV [5]. It is thought to have an animal (zoonotic) origin, and is 96% identical at the whole genome level to other bat coronavirus samples (BatCov RaTG13) [6]. This is the seventh known coronavirus to infect people [7]. The coronaviruses are capable of causing illnesses ranging from the common cold to more severe diseases such as Middle East respiratory syndrome (MERS, fatality rate ∼34%). The standard method of testing is polymerase chain reaction (PCR) [8]. Based on Johns Hopkins University statistics, the global Case fatality rate (CFR) is 2.36 % as of 24 November 2020 [3]. CFR strongly varies by region – from less than 1% in Norway, Iceland, Slovakia, Cyprus, Israel, Kuwait, Sri Lanka, and Malaysia to 5-10% in Mexico, Ecuador, Bolivia, Egypt, China, and Iran [9].

Several factors have been found statistically that increase severe course and mortality of COVID-19:

1. One of the most important factors in COVID-19 mortality is the age of infected. The Case fatality rate (CFR) is the number of deaths divided by the number of diagnosed cases. According to data for China, CFR remains within 0.2% for children and young people in the age groups 0-9, 10-19, 20-29 and 30-39 years. With age, CFR increases rapidly and reaches 14.8% for the age group 80+ years [10].
2. The next crucial factor of mortality from COVID-19 appears comorbidity of patients with severe diseases. Most of those who die of COVID-19 have pre-existing (underlying) conditions, including hypertension, diabetes mellitus, and cardiovascular disease [11, 12].
3. Vitamin D deficiency in populations. Strong correlation between prevalence of severe Vitamin D deficiency and population mortality rate from COVID-19 in Europe has been found in [13]
4. Very high Body mass index (Obesity). Johns Hopkins cardiologist David Kass discusses a recent study he co-led that links higher body mass index to more severe cases of COVID-19 and points to obesity as a significant pre-existing condition in younger patients in particular [14].
5. The role of the BCG vaccine for the prevention of COVID 19 appears insignificant [15, 16].

The Infection fatality rate (IFR) reflects the per cent of infected individuals (diagnosed and undiagnosed) who die from a disease. IFR at least is several times less than CFR because of many asymptomatic and mild symptoms infected persons. It is hardly be accurately calculated yet the global value is of the order of 0.64% [17]. IFR strongly varies across regions and countries – from 0.11 % in Sub-Saharan Africa to slightly above 1 % in Western Europe and High-income Asia Pacific [18].

The basic reproduction number (R0) of the virus has been estimated to be 5.7 [19]. Therefore each infection from the virus is expected to result in 5.7 new infections when no members of the community are immune and no preventive measures are taken. This shows that COVID-19 is much more contagious than seasonal flu, which has R0 between 1 and 1.5. Preventive measures to reduce the chances of infection include staying at home, avoiding crowded places, keeping distance from others, washing hands with soap and water often and for at least 20 seconds, avoiding touching the eyes, nose, or mouth with unwashed hands, and monitoring and self-isolation for people who suspect they are infected. Authorities worldwide have responded by implementing travel restrictions, lockdowns, workplace hazard controls, and facility closures. Many places have also worked to increase testing capacity and trace contacts of infected persons.

Many countries have recommended that healthy individuals wear face masks at least in certain public settings. This recommendation is meant to reduce the spread of the disease by asymptomatic and pre-symptomatic individuals and is a complementary measure to established preventive measures such as social distancing. The severity of COVID-19 varies. The disease may take a mild course with few or no symptoms.

As of 13 March 2020, the WHO considered Europe the active centre of the pandemic [20]. On 19 March 2020, Italy overtook China as the country with the most reported deaths [21]. By 26 March, the United States had overtaken China and Italy with the highest number of confirmed cases in the world [22]. Despite being the first area of the world hit by the outbreak, the early wide-scale response of some Asian states, particularly South Korea, Taiwan, and Vietnam, has allowed them to fare well [23, 24, 25].

After China in mid-March, the pandemic spread and caused high levels of infection in South Korea, Australia, Iran and Western Europe, especially in Italy, Spain, Iceland and Scandinavia. By mid-April, the pandemic had spread significantly to Turkey, Russia, Belarus, the United States, Canada and Saudi Arabia. By mid-May, the pandemic had spread significantly to South America, especially Brazil, Chile and Peru. In mid-June, the infection rate rose significantly in India, Pakistan, Afghanistan, Kazakhstan, South Africa and Mexico [9, 26].

The Fig. 1 shows a world map of countries by Cases per million people [27].

**Fig. 1.**
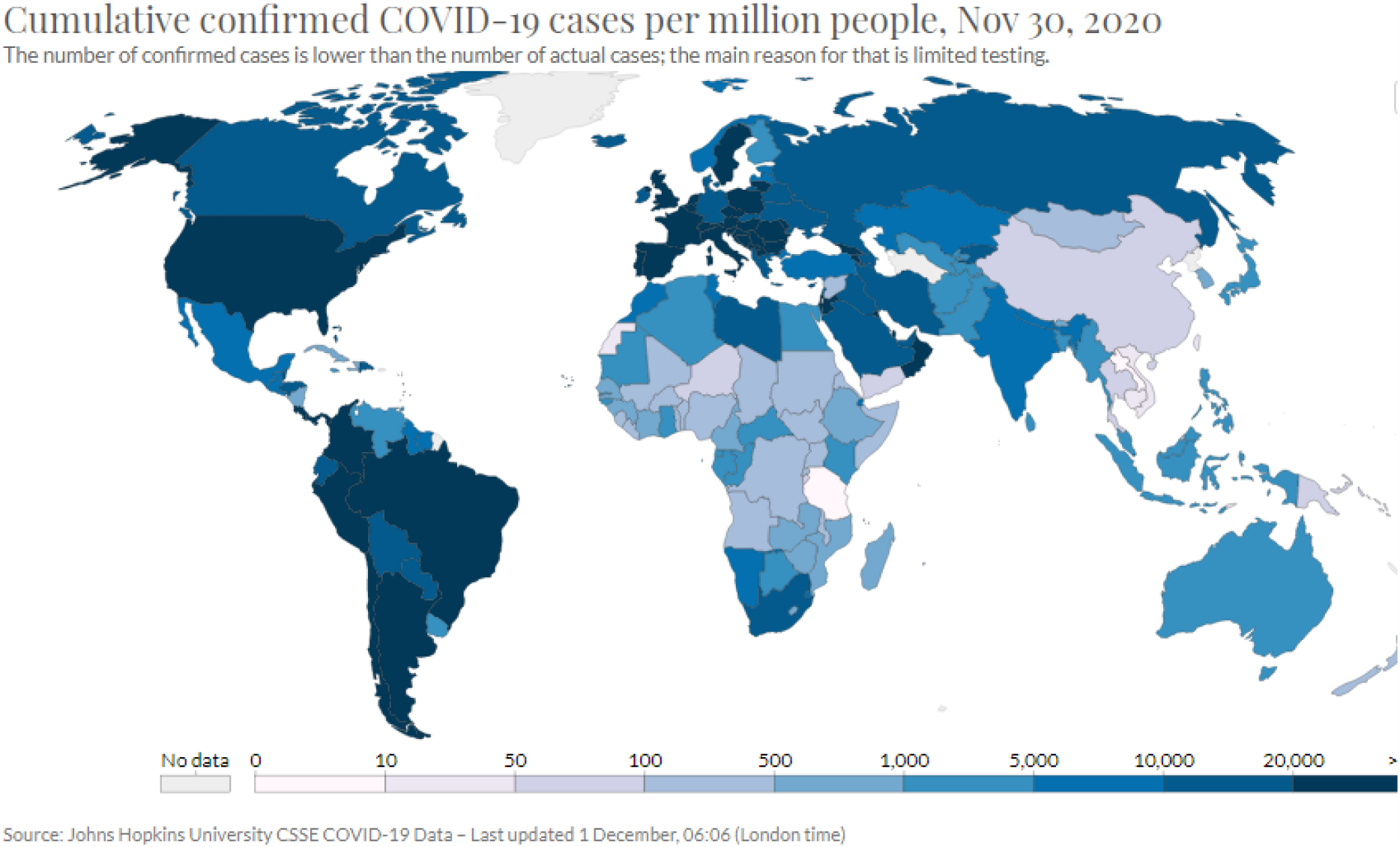
World map of countries by COVID-19 Cases per million people.

The Fig. 2 shows a world map of countries by COVID-19 Deaths per million people [28].

**Fig. 2.**
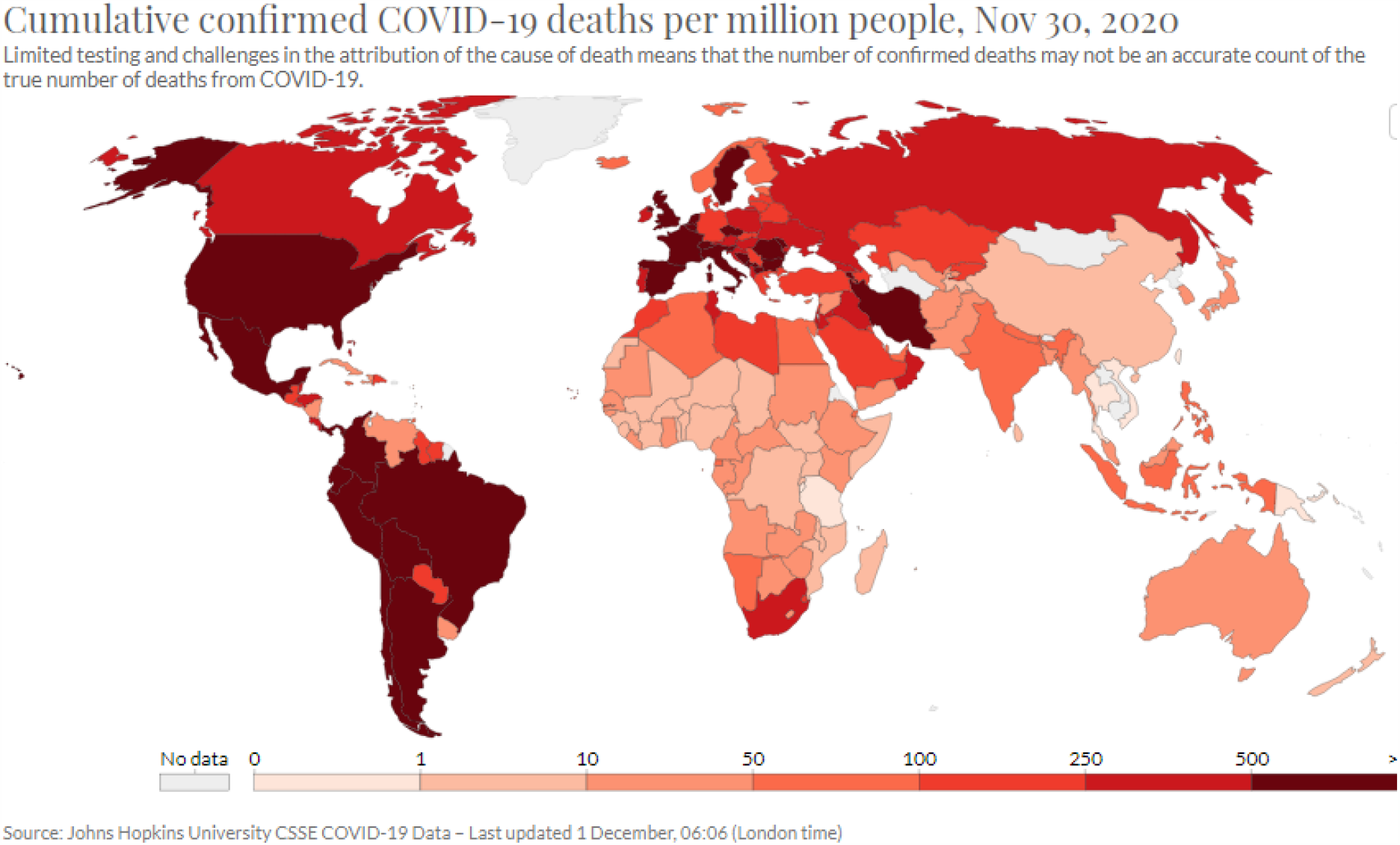
World map of countries by COVID-19 Deaths per million people.

## II. DATA AND METHODS OF PROCESSING

We used data for countries with a population of over 30 million. There are 45 countries from China with 1434 million inhabitants to Angola with 31.8 million inhabitants [29].

In our work, we have used the limit of more than 30 million populations, because these countries are less than a quarter of the countries in the world, and 85.8% of the world’s population lives in them. Moreover, the difference in population of the countries used for statistical analysis does not exceed 50 times. If all countries were used in the study, taking into account that the difference in the population between the largest country and smallest one (China and the Vatican, respectively) is over a million times, an unacceptably large difference in the statistical weight of the countries would be obtained.

Data for total COVID-19 cases per 1 million population (Cases/1M) and deaths per 1 million populations (Deaths/1M) were used for all countries studied. The data were taken from [9]. Data for 28 May 2020 were used in the main calculations. To check whether there is a change in the correlations with the development of the pandemic, a statistical analysis was made for four different dates – 9 April, 28 May, 7 August and 30 November 2020.

The following 10 demographic, economic and social indicators (indices) for the surveyed countries were used, namely:

1. Life expectancy at birth (Life Expectancy) [30].
2. Median age [31].
3. Population growth rate (Growth Rate) [32].
4. Population Density [33].
5. Gross domestic product at purchasing power parity per capita – GDP (PPP) [34].
6. Human Development Index (HDI) [35].
7. Gini index of income equality (Gini) [36].
8. Intelligence Quotient (IQ) of countries [37].
9. Corruption Perceptions Index (CPI) [38].
10. Democracy Index [39].

In addition to linear regression, power, logarithmic and exponential functions were used for statistical data processing. The statistical significance of the found correlations was assessed using Student’s t-test. The statistical package STATISTICA has been used for calculations.

## III. RESULTS

Below the results of statistical studies on the relationship of total COVID-19 cases per 1 million population and deaths per 1 million populations at 28 May 2020 with 10 demographic, economic and social indicators (indices) are shown. We have found that the Log Deaths per million is the most closely connected with Log Cases per million and the coefficient of correlation reaches R = 0.926. The dependence of Log Deaths per million from Log Cases per million is shown in Fig. 3. The names of the most deviated countries from the regression line are shown in the figure.

**Fig. 3.**
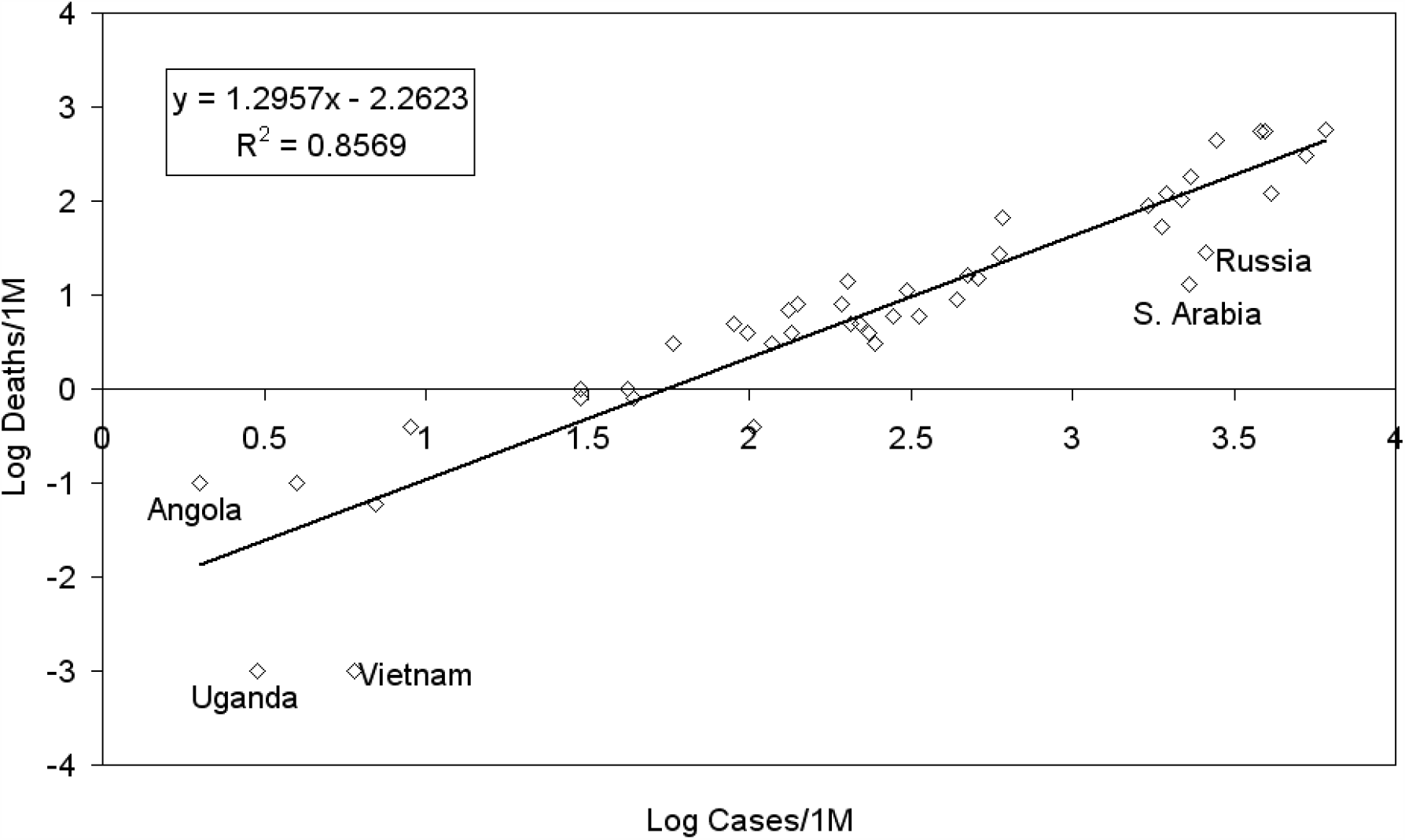
Dependence of Deaths per million from Cases per million.

Therefore, it is statistically clear that how the more are Cases per million in a country, the more are Deaths per million. From Fig. 3 it is seen that Saudi Arabia, Russia, Vietnam and Uganda lie below the regression line, i.e. mortality in these countries is lower.

A statistical analysis of the relationship of total COVID-19 cases and Deaths with 10 demographic, economic and social indices has been implemented below.

A statistical link between total Cases and Deaths with Life Expectancy was first sought. A positive correlation was found with correlation coefficients R = 0.532 and R = 0.514, respectively. As total Cases and Deaths per 1 million differ by several orders of magnitude in different countries, we hypothesized that they increase exponentially with Life Expectancy. Indeed, the correlation coefficients of the exponential relationship increased and reached 0.634 and 0.594, respectively. The dependence of Log Cases/1M from Life Expectancy is shown in Fig 4. The most deviating countries from exponential relationship were shown. Vietnam, Japan, South Korea, Thailand, Myanmar and Angola lie below the regression line, whereas Spain, USA, Peru, Russia, South Africa, Nigeria and DR Congo lie above the regression line.

**Fig. 4.**
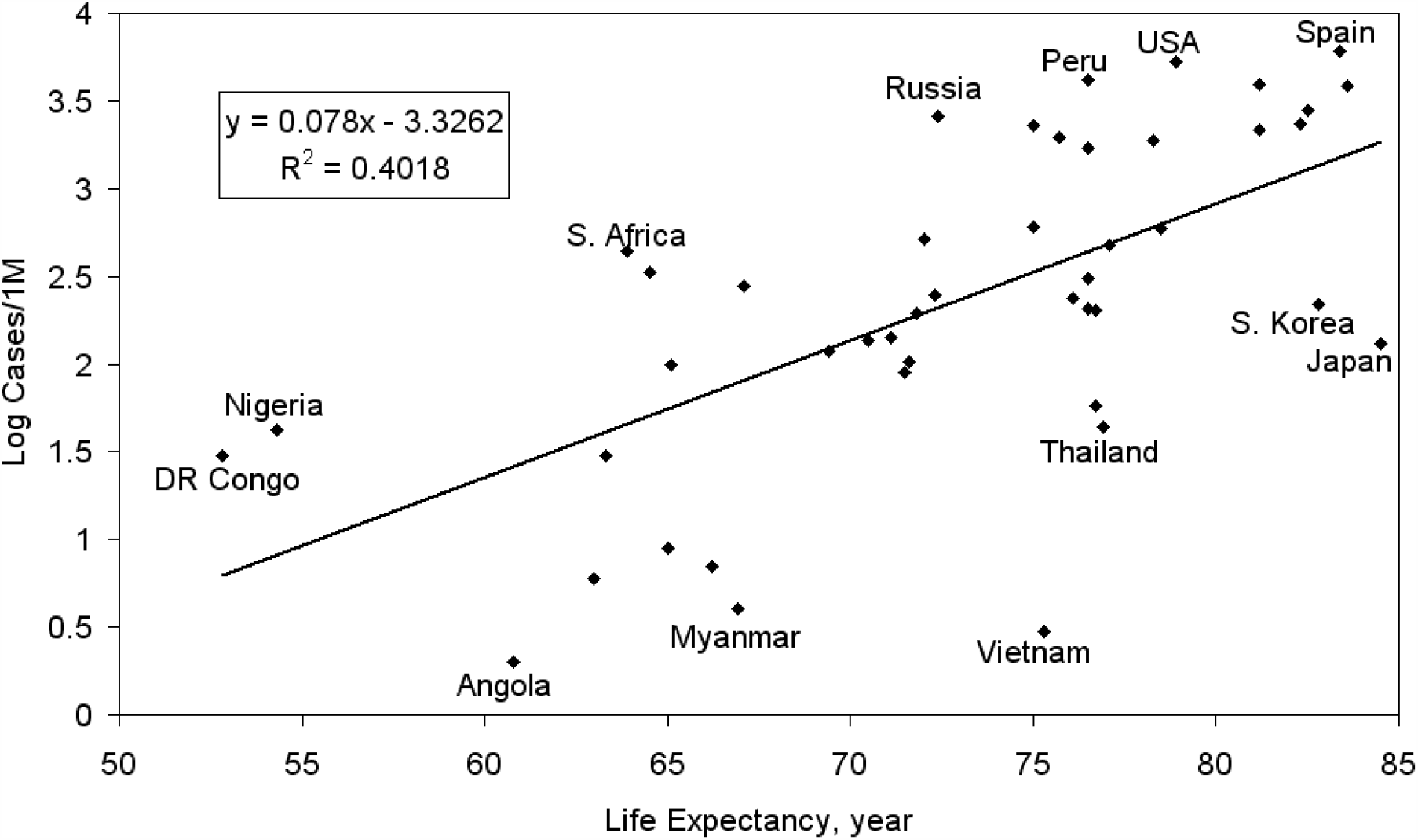
Dependence of Cases per million from Life Expectancy.

The statistical significance of the found correlations determined using Student’s t-test was p <0.0001. The trend of exponential growth of total Cases and Deaths with Life Expectancy is unambiguous. This is not a surprising result as it is well known that COVID-19 disease affects people over the age of 60 much more severely [10]. Due to the high proportion of adults over the age of 65 living in countries with a high Life Expectancy, their Cases and Deaths are significantly higher than those with low Life Expectancy.

The Median Age of a population is the point at which half the population is older than that age and half is younger. The Median Age closely correlated with Life Expectancy (R = 0.845). By reason of that the correlations of Log Cases and Log Deaths with Median Age were similar to Log Cases and Log Deaths with the Life Expectancy, respectively R = 0.613 and R = 0.596. The relationship of Log Deaths per million from Median Age is shown in Fig. 5. The countries that deviate significantly from the exponential curve are also shown.

**Fig. 5.**
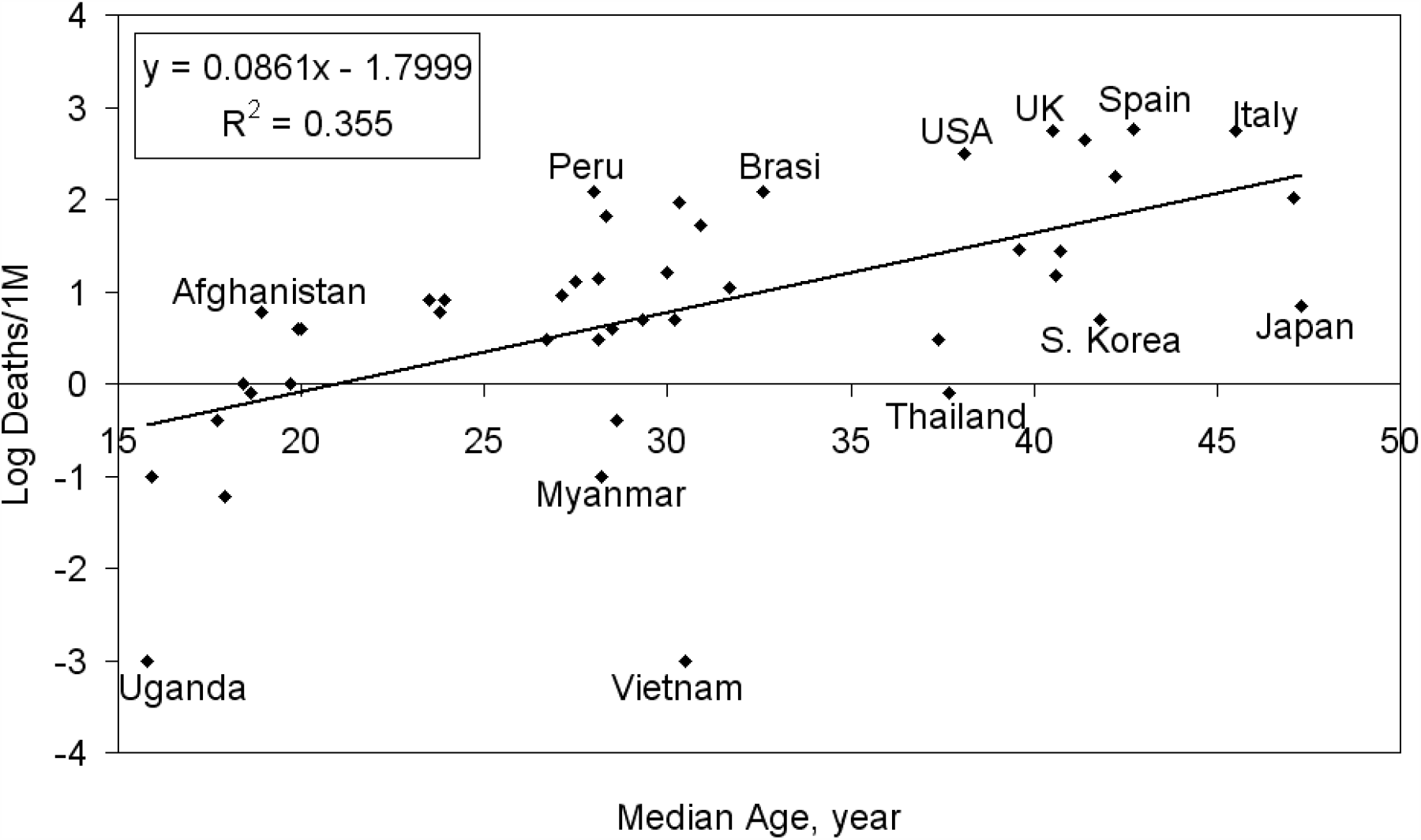
Dependence of Deaths per million from Median Age.

The next studied demographic indicator is population Growth Rate. There is a close negative correlation between Growth Rate and Life Expectancy (R = - 0.775) and because of this the Log Cases and Log Deaths show negative correlations R = - 0.540 and R = - 0.531 with the Growth Rate, respectively.

The last studied demographic indicator is the Population Density. The usual expectations are that the countries with higher Population Density will have higher total Cases and Deaths per 1 million. The statistical analysis has shown a surprising result - absence of significant correlation of Cases and Deaths with Population Density. The found negative coefficients of correlation of Cases and Deaths are: R = - 0.203 and R = - 0.20, respectively. These correlations are low and not statistically significant, therefore Cases and Deaths don’t depend from the Population Density.

The correlations of Cases and Deaths per 1 million was then sought with a purely economic index, namely Gross domestic product at purchasing power parity per capita (GDP). Close correlation of Log Cases and Log Deaths with Log GDP was established having correlation coefficients R = 0.687 and R = 0.660, respectively. The results for Cases are presented in Fig. 6.

**Fig. 6.**
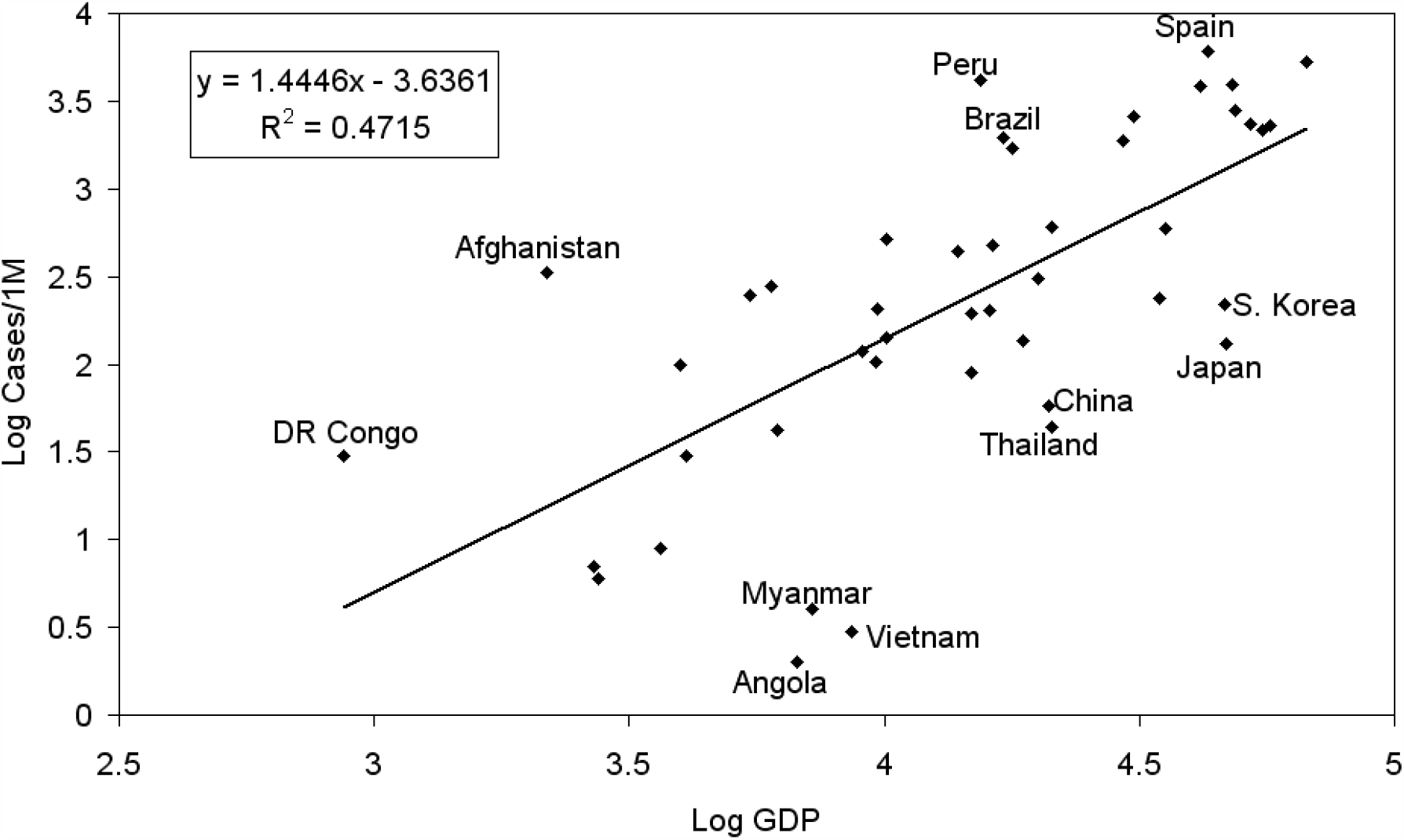
Dependence of Cases per million from GDP (PPP) per capita.

The countries that deviate most significantly from power law are shown on the figure. Some of countries below the regression line have a few Cases because of strict quarantine for example Japan, South Korea, Vietnam, and China. The countries above the regression line as Spain, Peru and Brazil has exceedingly cases because of late and/or insufficient effective pandemic restrictions.

Therefore, the total Cases and Deaths per 1 million increase by degree law with Gross domestic product per capita. This result is astounding because it shows that the higher the Gross domestic product per capita, the higher the rate of infection and mortality from COVID-19.

The next index included in statistical calculations is Human Development Index (HDI). The HDI is a statistic composite index of life expectancy, education, and per capita income indicators, which are used to rank countries by tiers of human development. It is accepted that HDI evaluates development not only by economic advances but also improvements in human well-being. The dependence of Log Cases and Deaths per million from HDI is shown in Fig. 7. The Cases per million are shown in blue and the Deaths per million are in red.

**Fig. 7.**
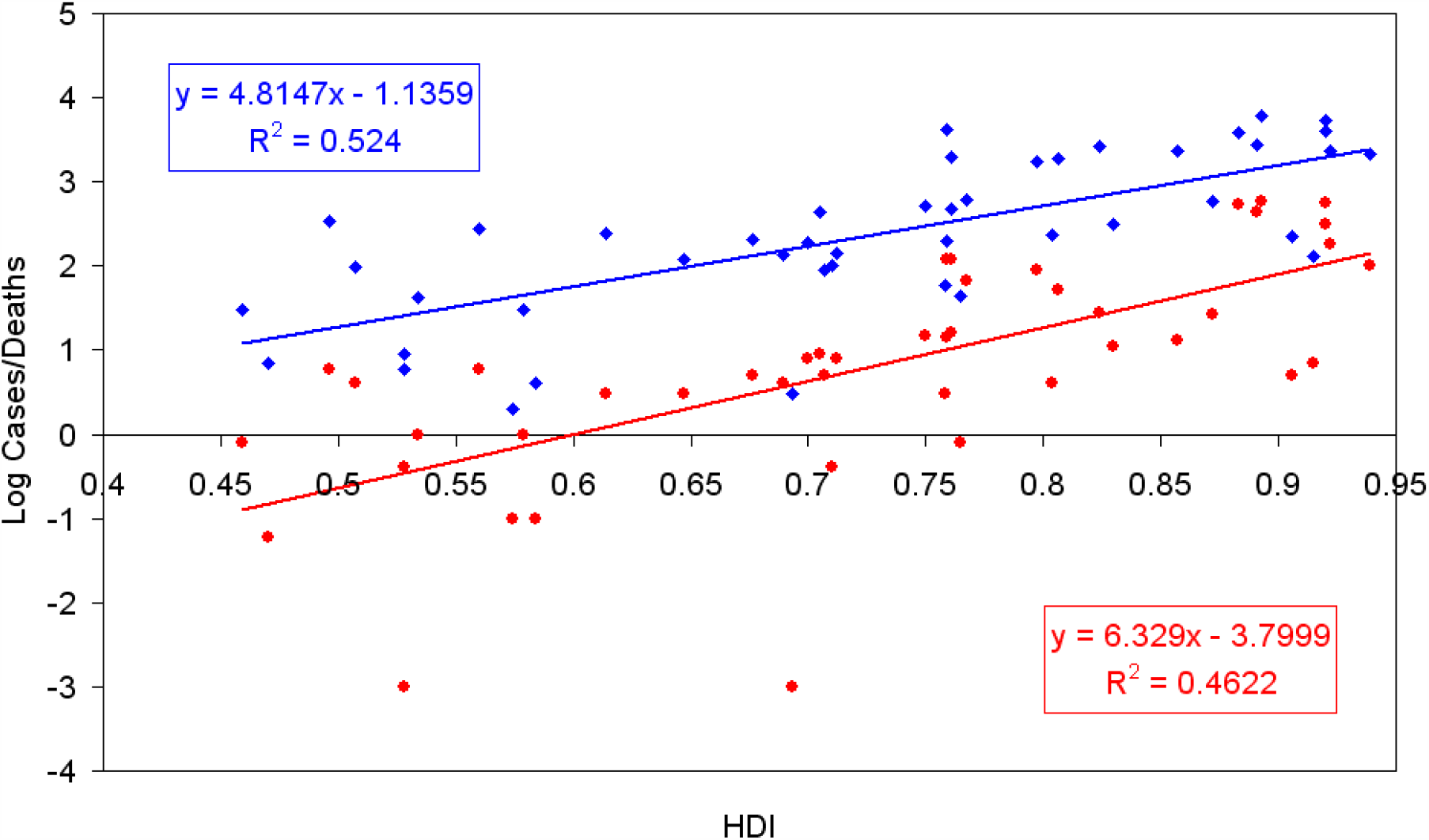
Dependence of Cases and Deaths per million from Human Development Index.

The correlation coefficients reach high values of R = 0.724 and R = 0.680, respectively. This result confirms above mention observation that the richest and well-being countries are most severe affected from the COVID-19 pandemic. This paradoxical result cannot be simply explained, because countries with high per capita incomes, well-developed health systems and a high level of education should be expected to be better protected from a pandemic. Apparently, in the case of the COVID-19 pandemic, the population of these countries is significantly more vulnerable than countries with a lower Human Development Index. The main reason for this is that the population in countries with high HDI is characterized by high mobility and is not prone to sufficient pandemic restrictions, with the exception of some countries in East Asia.

Geographic distribution of countries according Human Development Index [43] is presented in Fig. 8. It can be seen that this distribution is very similar to the distribution of countries by COVID-19 Deaths per million people presented in Fig. 2.

**Fig. 8.**
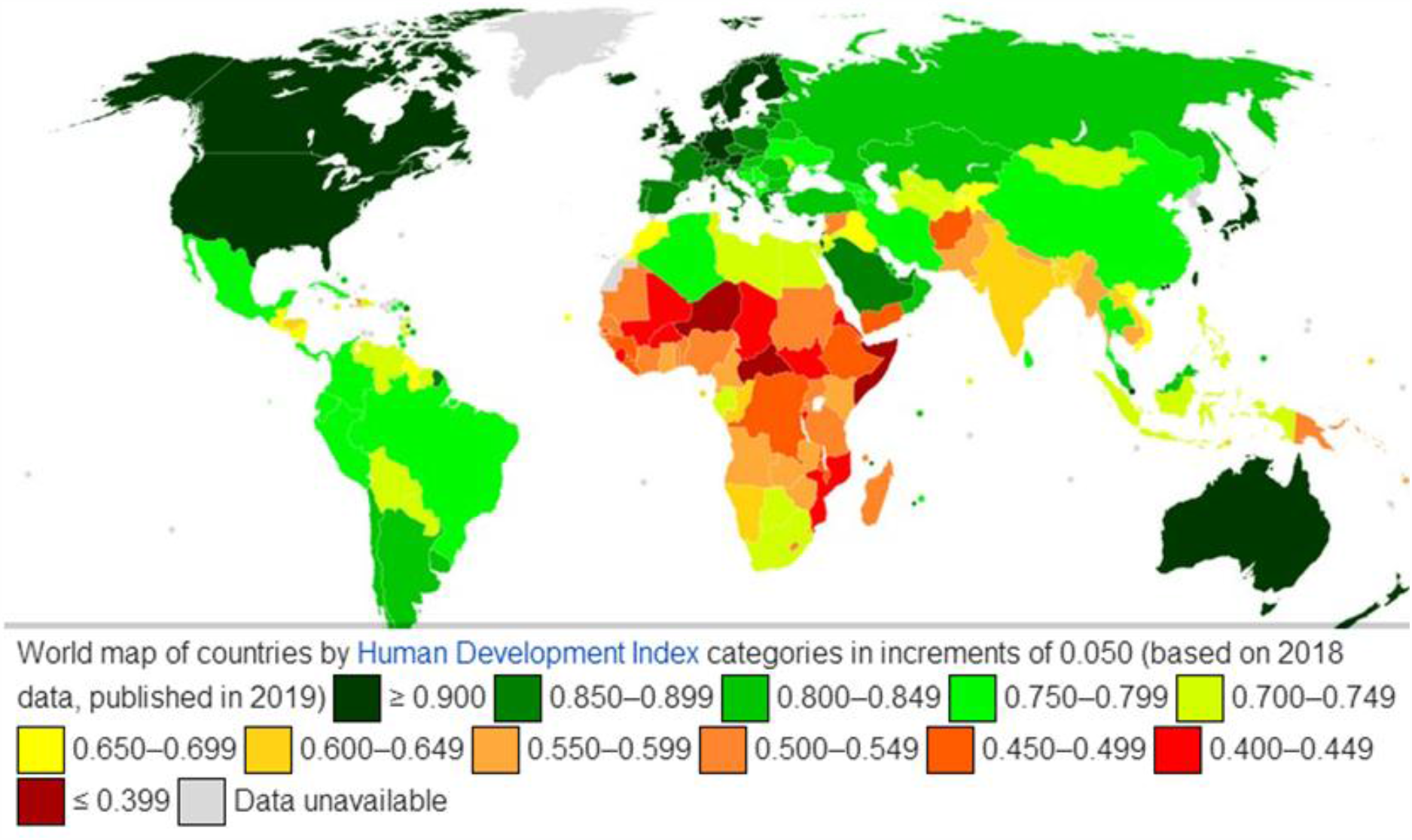
World map of countries by Human Development Index.

It has been found that the total Cases and Deaths per million showed no correlations with Gini index of income equality. Therefore, Cases and Deaths don’t depend from income equality.

The statistical relationship of Log Cases and Deaths per million with Corruption Perceptions Index (CPI) has been analysed, too. The found correlation coefficients are R = 0.534 and R = 0.542, respectively. The Corruption Perceptions Index is closely related with HDI (R = 0.785) and the last index could explain the above correlation.

The situation is similar with the connection of Log Cases and Deaths per million with Intelligence Quotient (IQ) of countries. The respective correlation coefficients are R = 0.487 and R = 0.432, respectively. The IQ of countries is also closely correlated with HDI (R = 0.80). Therefore, the defining parameter is again the Human Development Index.

Finally, we found correlation of Log Cases and Deaths per million with Democracy Index R = 0.459 and R = 0.504, respectively. The Democracy Index is also well correlated with HDI (R = 0.616).

The found statistically significant relationships of COVID-19 cases and Deaths with most of studied demographic, economic and social indicators are the result of their connections with Log GDP PPP. The correlation coefficients *R* and significance levels *p* of the connection of Log GDP PPP with the rest indicators is presented in Table 1.

**Table 1.** Correlation coefficients and significance levels of the relationships of Log GDP PPP with the rest indicators.

|  | Correlation coefficient | Significance level |
| --- | --- | --- |
| Human Development Index | 0.962 | 0.00001 |
| Life Expectancy | 0.858 | 0.00001 |
| Median Age | 0.810 | 0.00001 |
| Growth Rate | - 0.728 | 0.00001 |
| Intelligence Quotient | 0.746 | 0.00001 |
| Corruption Perceptions Index | 0.756 | 0.00001 |
| Democracy Index | 0.577 | 0.0001 |
| Gini index of income equality | -.0192 | n. s. |
| Population Density | -0.050 | n. s. |

Table 1 shows that the studied indicators excluding Density of population and Gini index have statistically significant correlations with Log GDP PPP.

To check whether there is a change in the correlations with the development of the pandemic, a statistical analysis was made for four different dates – 9 April, 28 May, 7 August and 30 November 2020. The dynamics of the correlation coefficient R between Cases and Deaths, on the one hand, and HDI, Life Expectancy, IQ, Growth Rate and Population Density, on the other hand, are shown in Figure 9. Significance level was shown by dotted lines at levels p = 0.05 and p = 0.001.

**Fig. 9.**
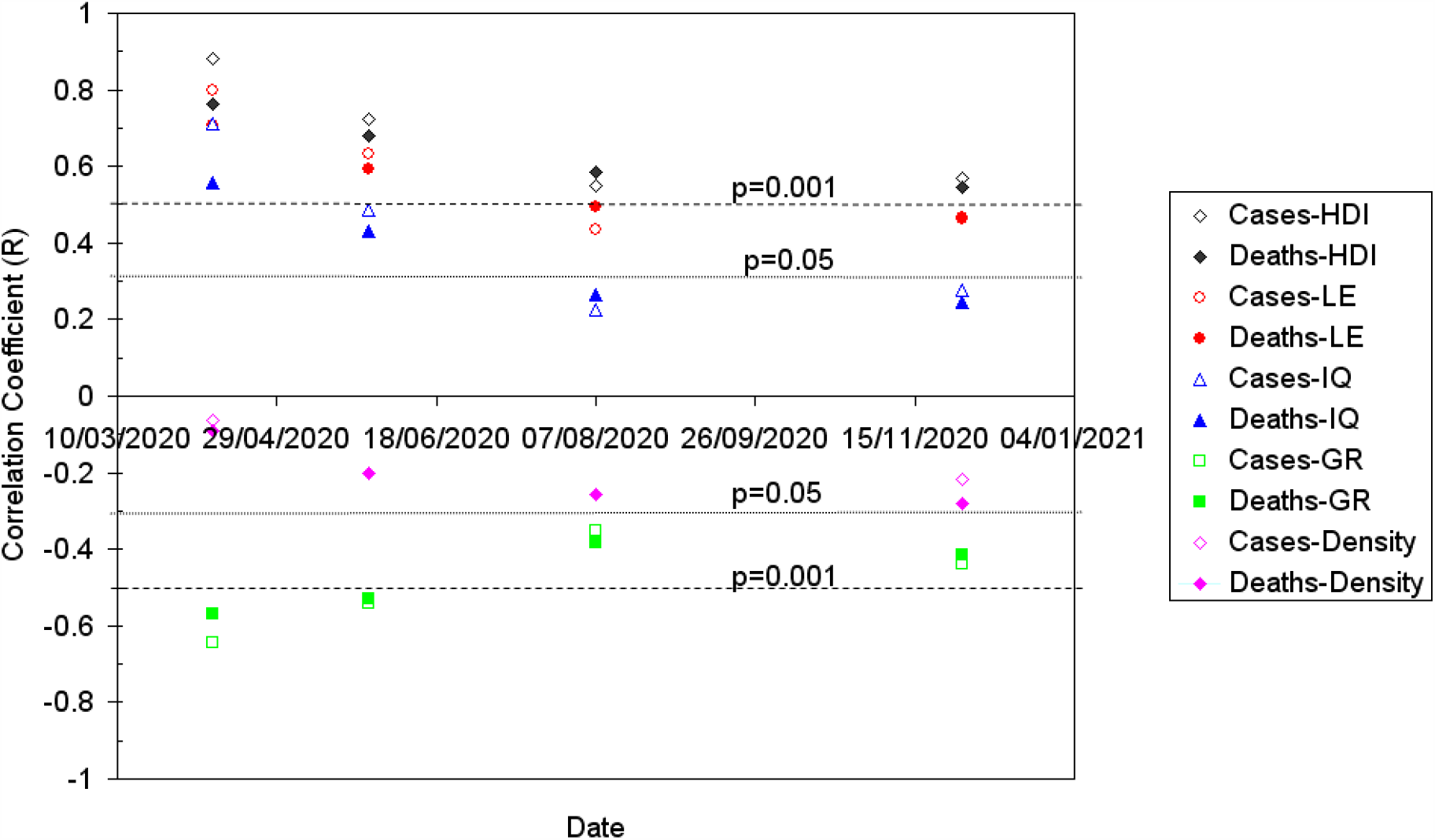
The dynamics of the correlation coefficient R between Cases and Deaths, on the one hand, and HDI, Life Expectancy, IQ, Growth Rate and Population Density, on the other hand.

Fig. 9 shows that the correlation coefficients of COVID-19 cases and Deaths with the rest indicators decrease during the pandemic. A probable reason for this time course of correlation coefficients is that most West European countries with high GDP, HDI, and Life Expectancy (Italy, Spain, UK, France, Germany), Canada and USA introduced stricter pandemic restrictions than at the beginning of the pandemic and they succeeded significantly slow the spread of the infection. On the other hand, in recent months, the pandemic has spread rapidly to a number of countries with medium or low GDP, HDI, and Life Expectancy in the Latin America (Mexico, Brazil, Argentina, Peru, Colombia), India and South Africa. In these countries, the pandemic began later, and when the number of COVID-19 cases and Deaths in Western Europe and the United States increased alarmingly, the Cases and Deaths in the indicated countries remained low. This created illusions among the people and institutions in these countries that the pandemic would not affect them severely, and therefore pandemic restrictions were significantly weakened. As a result, COVID-19 cases and Deaths have risen sharply in these countries. This confirms that in case of dangerous infectious diseases against which there is no specific medication or mass vaccination, the only effective means of limiting the percentage of infected are pandemic restrictions. For this reason, the latter play a crucial role in limiting the infection rate and the associated mortality. The main thing is the discipline and self-discipline of the population, its readiness to comply with pandemic restrictions especially social distance, isolation and face masks.

## IV. DISCUSSIONS AND CONCLUSIONS

The statistical relationships of total COVID-19 Cases and Deaths per million populations in 45 countries, where 85.8% of the world’s population lives with 10 demographic, economic and social indicators were studied. The relationship of Deaths per million population and total Cases per million population is very close and reaches correlation coefficient R = 0.926. It is interesting that the close correlations were found of Cases and Deaths per 1 million with a purely economic index like GDP PPP per capita, where R = 0.687 and R = 0.660, respectively. Even more close correlations were found of Cases and Deaths per 1 million with a composite index HDI, where the correlation coefficients reach 0.724 and 0.680, respectively. The main reason for these paradoxical results is the underestimation of pandemic restrictions in the form of masks, social distance and disinfection in most of these countries. Other indicators (excluding Gini index and Population Density) also show statistically significant correlations with Cases and Deaths per 1 million with correlation coefficients from 0.432 to 0.634. The statistical significance of the found correlations determined using Student’s t-test was p <0.0001. Surprisingly, there was no statistically significant correlation between Cases and Deaths with Population Density.

The established close positive correlation between Cases and Deaths per million with GDP (PPP) per capita and HDI seems paradoxical at first sight. Because countries with high GDP (PPP) per capita and HDI have well-developed health systems that are provided with sufficient high-tech medical equipment and highly qualified specialists. What, then, is the reason why most of these countries allow the highest rates of COVID-19 infection and mortality? These countries do have well-off and well-functioning health systems in a normal health situation. In this case, however, the situation is not normal, but a crisis one, because we have a pandemic due to a new coronavirus SARS-CoV-2 against which there is no vaccine or specific medicine.

This coronavirus causes airborne infection and has high reproductive number and is very contagious. The situation is further complicated by the fact that the latency period of COVID-19 is too long and reaches 1-2 weeks. In addition, most cases of COVID-19 infection have mild flu-like symptoms or asymptomatic. Therefore, it is not possible to distinguish well between the sick and to isolate, and for the healthy to continue to work and live unrestricted by pandemic restrictions. In case of dangerous infectious diseases against which there is no vaccine, the only effective means of limiting the percentage of infected are pandemic restrictions. For this reason, the latter play a crucial role in limiting the infection rate and the associated mortality. Developing countries have significantly lower Median Age, obesity and levels of comorbidities of population than high-income countries. Therefore, the material well-being of the citizens and the provision of the health systems of the countries with equipment, consumables and qualified specialists play a secondary role. The main thing is the discipline and self-discipline of the population, its readiness to comply with pandemic restrictions especially social distance, isolation and face masks. This is why many countries with relatively low GDP per capita and HDI (Vietnam, Philippines, Ethiopia, Nigeria) have been able to achieve much lower COVID-19 infection and mortality than countries with high GDP per capita and HDI (Italy, Spain, USA, United Kingdom, France). Most countries with high GDP per capita and HDI are developed democracies with high individual freedom from the state and its organs, and their governments hesitated too long before introducing pandemic restrictions restricting the freedom of movement, assembly and life of citizens. Of course, there are countries with high GDP per capita and HDI, such as Japan, South Korea and Malaysia, which have managed to effectively limit the spread of COVID-19 and prevent high mortality. This is due to the traditional culture of these countries, in which the trust of the citizens in the ruling elite and their tendency to obey the reasonably justified decisions of these elite is deeply rooted.

## Data Availability

I accessed only publicly available data from the Web.

## ACKNOWLEDGMENTS

The author is very grateful to Prof V. Guineva for useful discussions.

## Author Declarations

No human subjects research as I accessed only publicly available data.

